# Investigating Neonatal Sepsis: Anti-Infectives, Diagnostics and Guidelines used in Health sysTems across Sub-Saharan Africa - The INSIGHTS Study

**DOI:** 10.1101/2025.11.05.25339619

**Authors:** Jack Stanley, David Hettle, Rachel Poffley, Larisse Bolton, Emelyne Gres, Isabel Coelho, Syeda Ra’ana Hussain, Gildas Boris Hedible, Gwendoline Chimhini, Valeriane Leroy, Madeleine Amorissani Folquet, Felicity Fitzgerald, Angela Dramowski

**Author notes:** shared first authorship (Jack Stanley, David Hettle). shared last authorship (Felicity Fitzgerald, Angela Dramowski).

## Abstract

**Background:** Sepsis is a leading cause of neonatal mortality in sub-Saharan Africa (SSA), where microbiological diagnostic capacity and antibiotic access are limited. High antimicrobial resistance (AMR) rates limit the effectiveness of current treatment guidelines, with concern that available antibiotics are rarely adequate treatment for neonatal sepsis in the region.

**Methods:** A cross-sectional online survey was electronically distributed in English, French and Portuguese to neonatal clinicians across SSA between April and June 2025. Questions focussed on the management of neonatal sepsis including diagnostic, antibiotic and guideline use. Responses were analysed descriptively and presented as percentages of the total number of responses.

**Results:** Of 169 responses (40/48 countries; 83.3%) from SSA, 71.6% were senior doctors, 88.8% managed neonatal sepsis at least weekly and 58.0% worked in central healthcare facilities. 9.0% of respondents never and 28.6% less than half of the time received blood culture results in time to impact patient care. Guidelines were almost universally used (97.6%). The commonest guideline for early-onset neonatal sepsis advised amoxicillin/ampicillin plus aminoglycosides (46.4% of responses). 50.3% of respondents had difficulties accessing antibiotics, with carbapenems and piperacillin-tazobactam least accessible. More than half of respondents (54.6%) had not attempted to author local guidelines, with insufficient local AMR data (45.6%) the most common barrier to guideline development.

**Conclusions:** This large survey highlighted widespread challenges in diagnostic and antibiotic access for neonatal sepsis in SSA. We find that clinicians rely on guidelines to guide starting antibiotics and to guide agent choice. Their practices reflect advice in global guidelines because attempts to author locally applicable guidelines are hindered by insufficient AMR data. These findings strengthen calls to improve microbiological diagnostic access and support data sharing to generate evidence-based, locally appropriate guidelines.

**Key Messages:** 

**What is already known on the topic:** Neonatal sepsis is a leading cause of death in sub-Saharan Africa. Amidst antimicrobial resistance (AMR) and structural barriers - limited diagnostic capacity, antibiotic access and few locally tailored guidelines, the management of sepsis is not well described, leaving a critical evidence gap.

**What this study adds:** This survey of neonatal clinicians in 40 of 48 countries in sub-Saharan Africa, is the most comprehensive to date. Timely diagnostic results are often unavailable, access to necessary antibiotics scarce, and clinicians rely heavily on guidelines, which are global directed rather than locally adapted, hampered by minimal AMR data.

**How this study might affect research, practice or policy:** Highlighting the importance of guidelines in clinicians’ practice must strengthen efforts to develop pan-African data sharing in neonatal sepsis, to enable locally relevant guidelines which can inform practice effectively amidst AMR and diagnostic, antibiotic and infrastructural challenges.

## Introduction

Sub-Saharan Africa (SSA) has the highest burden of neonatal mortality globally, with 27 deaths per 1000 live births[1], contributing to 46% of neonatal deaths globally[2] despite only accounting for 30% of live births[3]. Persistently high neonatal mortality is concerning against the backdrop of a global reduction in under five mortality and World Health Organization (WHO) Sustainable Development Goal (SDG) 3 targeting a reduction in neonatal mortality to 12 or fewer deaths per 1000 live births globally by 2030[1]. Recent estimates suggest that neonatal sepsis is responsible for 17-29% of neonatal deaths in SSA[2, 4]. Therefore, neonatal sepsis is a critical focus for reducing neonatal mortality and morbidity[5].

Effective management of neonatal sepsis requires prompt recognition, early administration of antibiotics and supportive care and timely microbiological diagnosis[6–8]. Diagnosing neonatal sepsis is challenging, as neonates present with non-specific clinical symptoms and signs[9]. A review of 14 SSA countries found that only 1.4% of medical laboratories provide bacteriological testing, an essential diagnostic for sepsis, recommended by the WHO[10], while a survey of medical practitioners across 25 SSA countries highlighted poor availability of blood culture and Gram staining[11]. Where microbiology services do exist, there are challenges of limited capacity, expertise, resources and training. Even when results are available, their ability to impact care is variable, affected by processing time, communication between microbiology and clinical teams and knowledge gaps in interpreting results[10]. For example, in Zimbabwe a study found that only 4% of blood culture results arrived in time to impact patient management[12]. With vast differences in service availability and quality within and between countries, the full picture of access to, and use of microbiology diagnostics in managing neonatal sepsis in SSA remains unclear.

Given these diagnostic challenges, current management guidelines rely on empirical antibiotic therapy while considering realistically available antibiotics[13]. Most health settings in SSA use WHO-based guidelines which have not been updated since 2005 and reflect international rather than local epidemiological patterns in antimicrobial resistance (AMR)[14]. Since then, antimicrobial susceptibility patterns have changed [13, 15] with SSA now having the highest AMR-attributable mortality globally[16]. There has been rapid development of AMR in the organisms commonly responsible for neonatal sepsis. Resistance to all WHO-recommended first-line antimicrobials amongst *Klebsiella* spp. increased from 7% to 68% between 2002 and 2017[17] and resistance also increased in this time to second-line therapy, which typically includes a third-generation cephalosporin[15, 18]. This correlates with high regional mortality caused by nosocomial neonatal sepsis due to *K. pneumoniae*, between 36%[19] and 71%[20].

There is growing opinion that these WHO guidelines may no longer be applicable for SSA amidst rising AMR[13]. Yet very few examples of national or hospital-level guidance informed by local data exist. Research to explore the reasons behind this remains limited. We surveyed clinicians caring for neonates across SSA to provide a comprehensive perspective on the current access to diagnostics and antibiotics and to better understand guideline utilisation in the management of neonatal sepsis in this region.

## Methods

### Study design

We created an online survey developed by a core group of six researchers and piloted for usability within the wider research team using REDCap[21, 22]. The survey comprised of 32 stem questions grouped into: demographics, microbiological diagnostic test availability, antibiotic use and availability and guideline utilisation. Most questions were closed-ended, with 5-point Likert scale type questions which addressed likelihood, frequency or agreement, and free-text responses for more nuanced information. The survey and accompanying information were available in English, French and Portuguese. The survey tool (English language version) is available in the supplementary material. Our target population was clinicians involved in the regular clinical care of neonates across 48 countries in SSA, defined as per World Bank classification[23]. An English language copy of the survey is attached as an appendix for reference.

We utilised a purposive and snowballing approach to sampling via the NeoNET AFRICA group, a multi-country, multi-disciplinary network of clinicians, researchers, epidemiologists and other specialists involved in neonatal sepsis across Africa[14] and the African Neonatal Association, a non-profit organisation representing over 250 African neonatologists and paediatricians working in neonatal care in SSA. We also distributed the survey via national and regional neonatal and paediatric networks (45 different organisations/networks contacted), and through social media and academic platforms to access relevant respondents.

All potential participants were provided with participation information on entry to the survey, after which consent was gained prior to survey response. Participants completed the survey anonymously and no financial incentives were offered. We aimed for responses from at least two-thirds of our 48 target countries, with a minimum of 100 respondents. The survey remained open for two months (03/04/2025-02/06/2025). Responses to most questions were compulsory, preventing closure of the survey without completion. In some instances, subsequent questions were displayed contingent on answers to earlier questions. In these instances, where the number of respondents might be less than the total number of participants, this is indicated in figure legends.

### Ethics

This study was approved by the Stellenbosch University Health Research Ethics Committee (HREC approval number: N24/10/118).

### Statistical analysis

Data was analysed in R version 4.4.2[24]. Answers to questions were reported as the percentage of those responding to the question. Some analyses were stratified by the respondent’s job role, place of work or the income status of the country they worked in according to the World Bank’s Income Groups[25] (low income, low-middle income, upper-middle income and high-income).

Categorical variables were expressed as percentages with the denominator being the total number of individuals providing responses to that individual question.

## Results

We received responses from 169 eligible respondents from 40 countries of the 48 targeted (no response from Equatorial Guinea, Eritrea, Eswatini, Guinea-Bissau, Mauritania, Seychelles, South Sudan, or Sudan). Three additional responses were excluded, two due to not being from a country targeted in the survey, and one due to inadequate details regarding country of practice and incomplete responses to several questions. Missing data was negligible due to the design of the survey, which required responses before progressing through the tool. The single episode mentioned above was early in the study and highlighted an issue which meant that all responses were not necessitated. This issue was thereafter resolved. Numbers of responses by country are shown in Figure 1. The countries with the most responses were South Africa (n = 25, 14.8%), Kenya (n=16, 9.5%), and Tanzania (n=11, 6.5%). The remaining 37 countries provided 1 to 9 responses.

**Figure 1.**
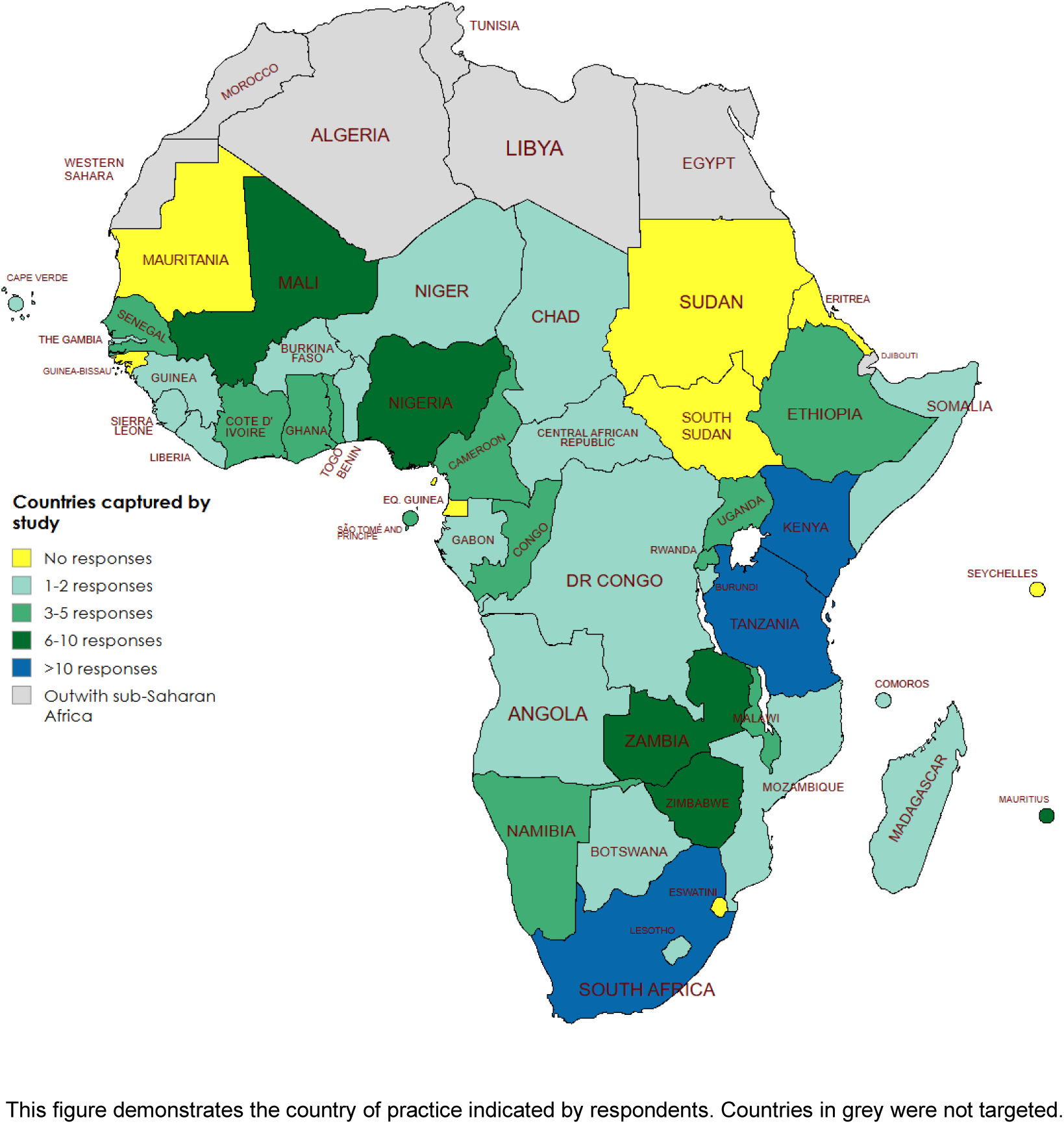
Location of Neonatal Sepsis Survey Respondents.

Responses were primarily from senior doctors (71.6%) (Figure 2). There was a spread in the size of the units in which respondents worked, with 31.4% working in units with less than 20 and 13.0% working in units with more than 100 neonatal inpatients on average. Most respondents (69.8%) worked in public facilities, with a further 19.5% working in both public and private facilities. The majority (58.0%) worked in central healthcare facilities with 27.8% working in district facilities and 15.4% in primary care.

**Figure 2.**
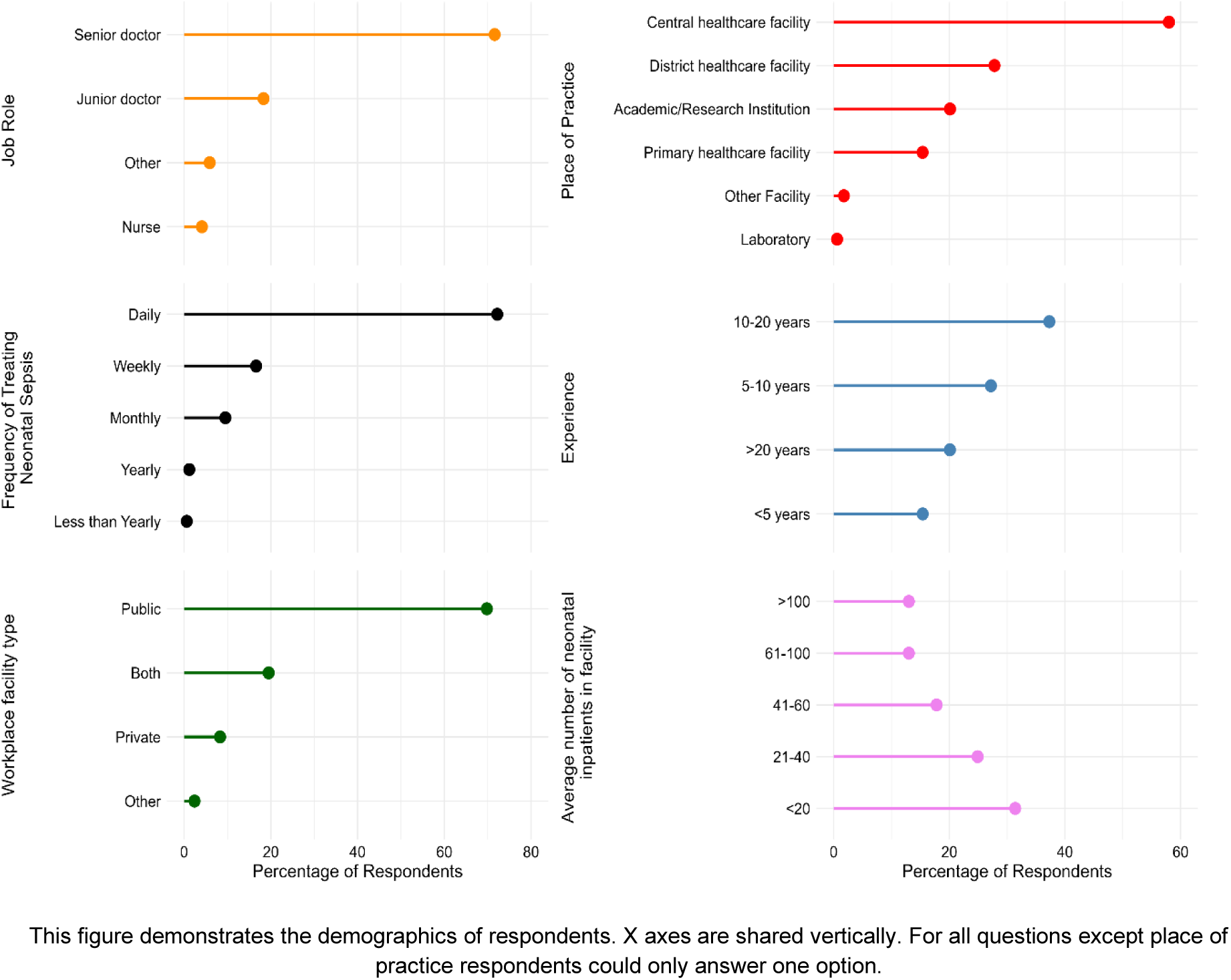
Demographics of Respondents.

Considering diagnostic investigations for suspected neonatal sepsis, white blood cell count was the test most commonly available in time to impact patient care (Figure 3), with 58.0% of respondents stating that this was always the case. 14.9% of respondents never and 28.6% less than half of the time received blood cultures in time to impact patient care, with all culture-based investigations available less frequently than biochemical tests. When the timely availability of tests are stratified by the income status of respondents’ country (Supplementary Figure 1), low- and low-middle income countries experience similar levels of difficulty accessing results in time to impact patient care, yet high-middle income countries have much better access to timely results for all tests. Of particular note, repondents from high-middle income countries rarely have difficulty obtaining timely blood culture results.

**Figure 3.**
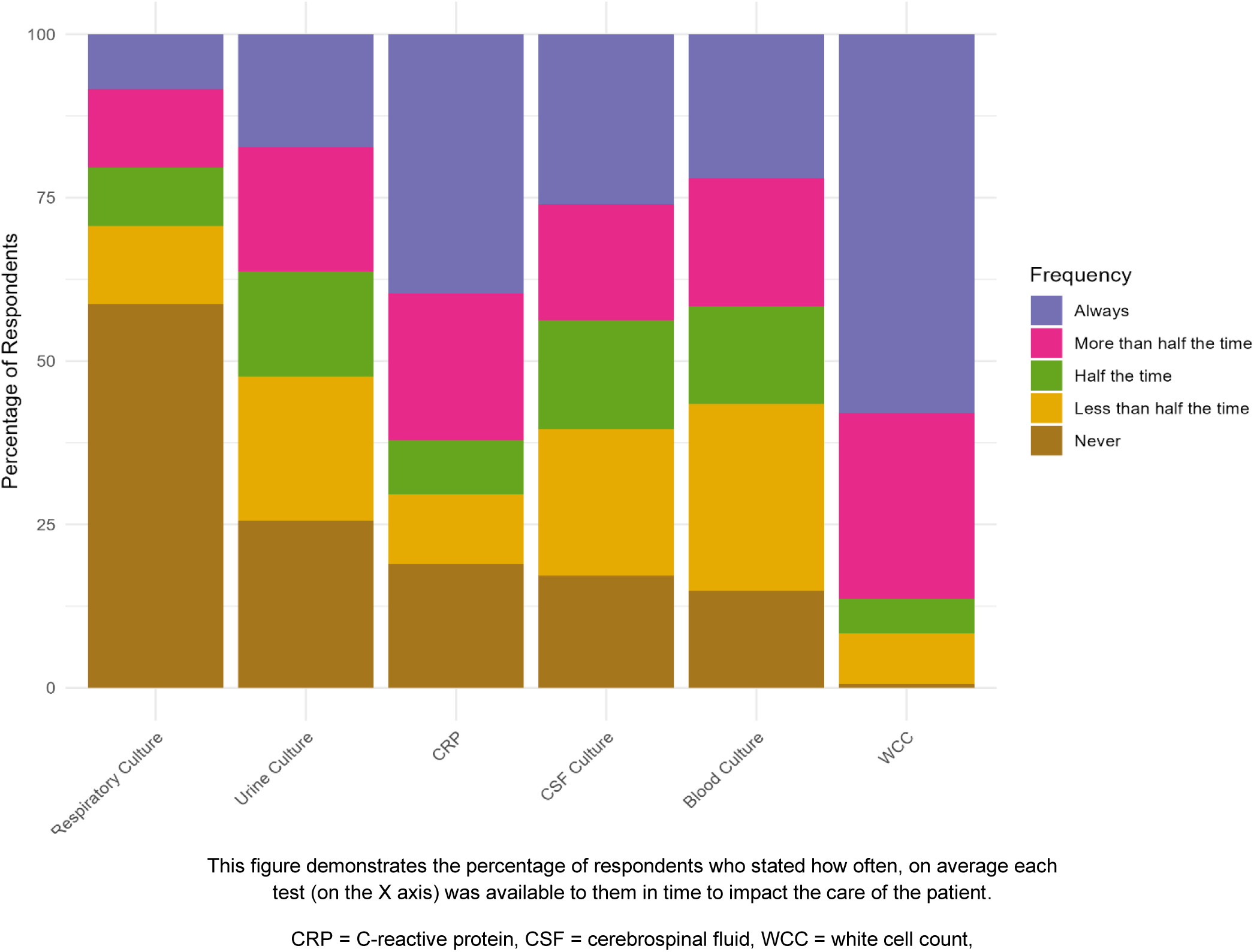
Test Result Availability in Time to Impact Patient Care.

The clinical condition of the neonate was the most important factor when deciding whether to start antibiotics (Figure 4), with most respondents reporting that they considered it always (69.6%) or more than half the time (22.4%). Guidelines were the next most commonly considered, with three-quarters of respondents considering them always (51.5%) or more than half the time (24.0%). On the other hand, families’ financial situation (69.6% of clinicians never influenced by this factor), family expectations (79.3% never), and colleague expectations (56.9% never) rarely influenced the decision to start antibiotics. There was little variation between senior doctors and other respondents (Supplementary Figure 2).

**Figure 4.**
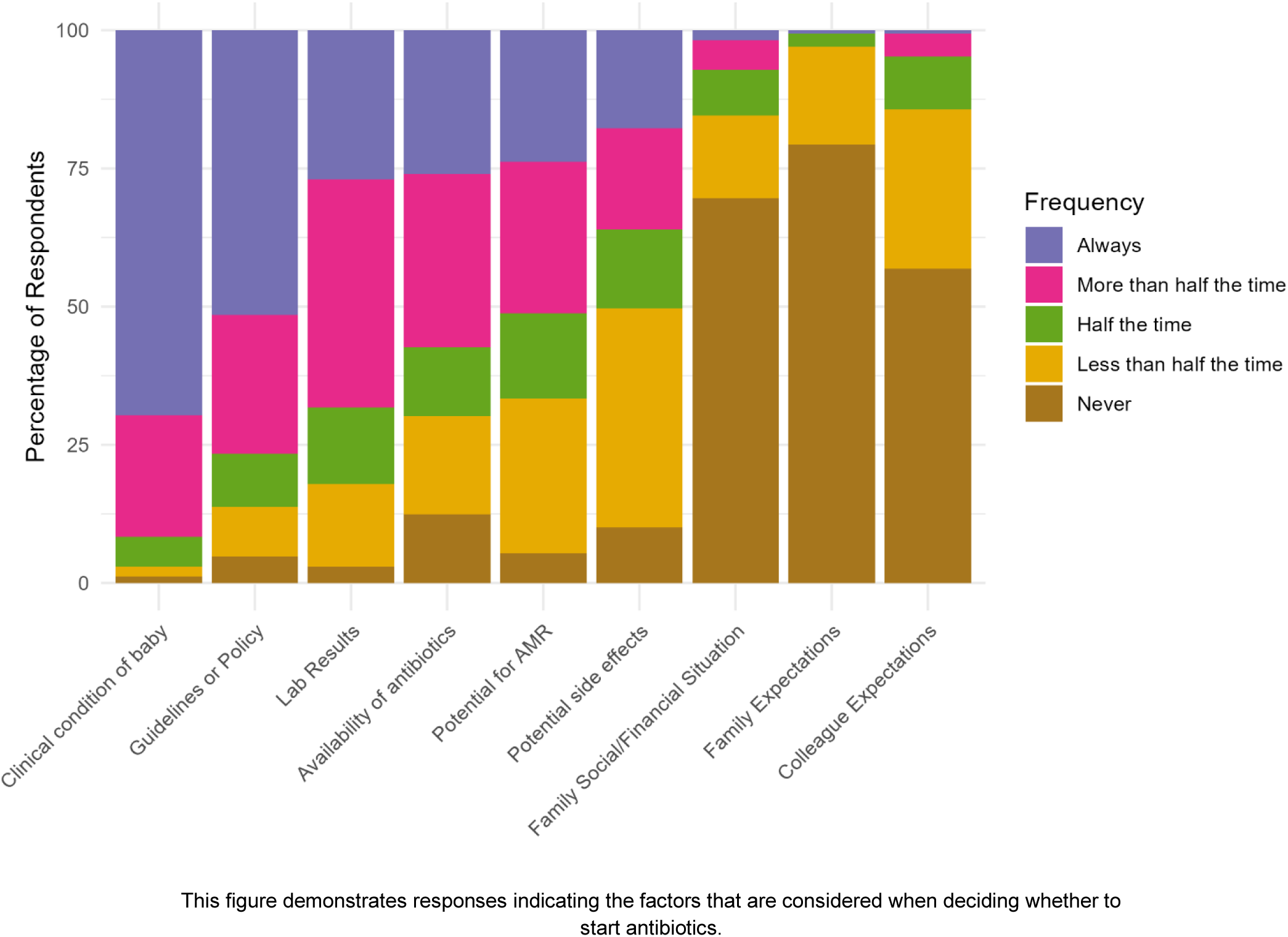
Influences on Decision to Start Antibiotics in Neonates.

When the decision has been made to start antibiotics, the choice of antibiotic agent was most influenced by the clinical condition of the baby (98.2%). Local (88.6%), national (81.4%) and international guidelines (78.2% of respondents) were used to direct choice of antibiotic agent.

Over half (50.3%) of respondents reported difficulty accessing antibiotics for treating neonatal sepsis. The most available antibiotics (Figure 5) were gentamicin (68.7% never had issues accessing), amoxicillin (64.1%), ampicillin (53.9%) and penicillins (49.3%). The most challenging antibiotics to access were colistin (52.9% always had difficulty accessing), piperacillin-tazobactam (38.1% always), vancomycin (25.6% always) and carbapenems (imipenem/meropenem, 23.4% always), all of which are classed as ‘Watch’ or ‘Reserve’ by the WHO AWaRe classification[26]. Supply was the most common factor limiting antibiotic access across all agents, most frequently reported for amikacin (48.4%), colistin (46.1%) and piperacillin-tazobactam (45.7%). Affordability is most frequently an issue for imipenem/meropenem (38.8%) and vancomycin (30.6%) (Supplementary Figure 3).

**Figure 5.**
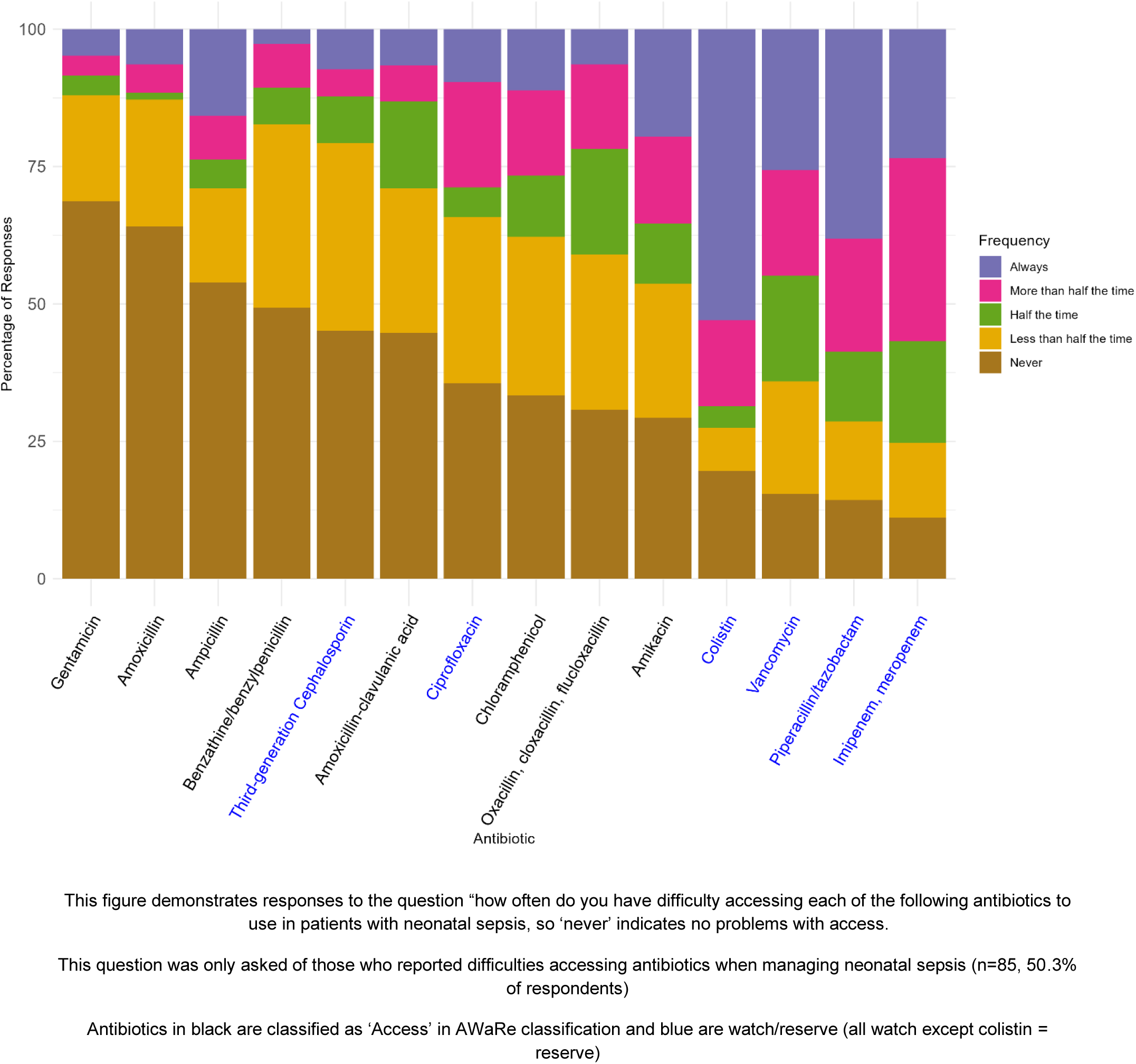
Difficulty Accessing Antibiotics for Treating Neonatal Sepsis.

Almost all respondents (97.6%) stated that they referred to guidelines (in any form) in the management of neonatal sepsis. 83% always and 15.6% sometimes considered these guidelines to be applicable to their setting.

Regarding the first-line antibiotic therapy for early-onset neonatal sepsis recommended by guidelines and used in practice (Figure 6), a combination of an aminopenicillin (amoxicillin or ampicillin) plus an aminoglycoside was most common (46.4% recommended in guidelines and 46.5% used in practice). Thereafter, penicillins (X-pen or benzylpenicillin) plus aminoglycoside (20.9% recommended in guidelines, 25.2% practice) and third-generation cephalosporin plus aminoglycoside (10.5% recommended in guidelines, 11.0% practice) were most common. In a minority of cases, monotherapy was suggested first-line. It is evident that what respondents use in practice closely mirrors what their guidelines recommend.

**Figure 6.**
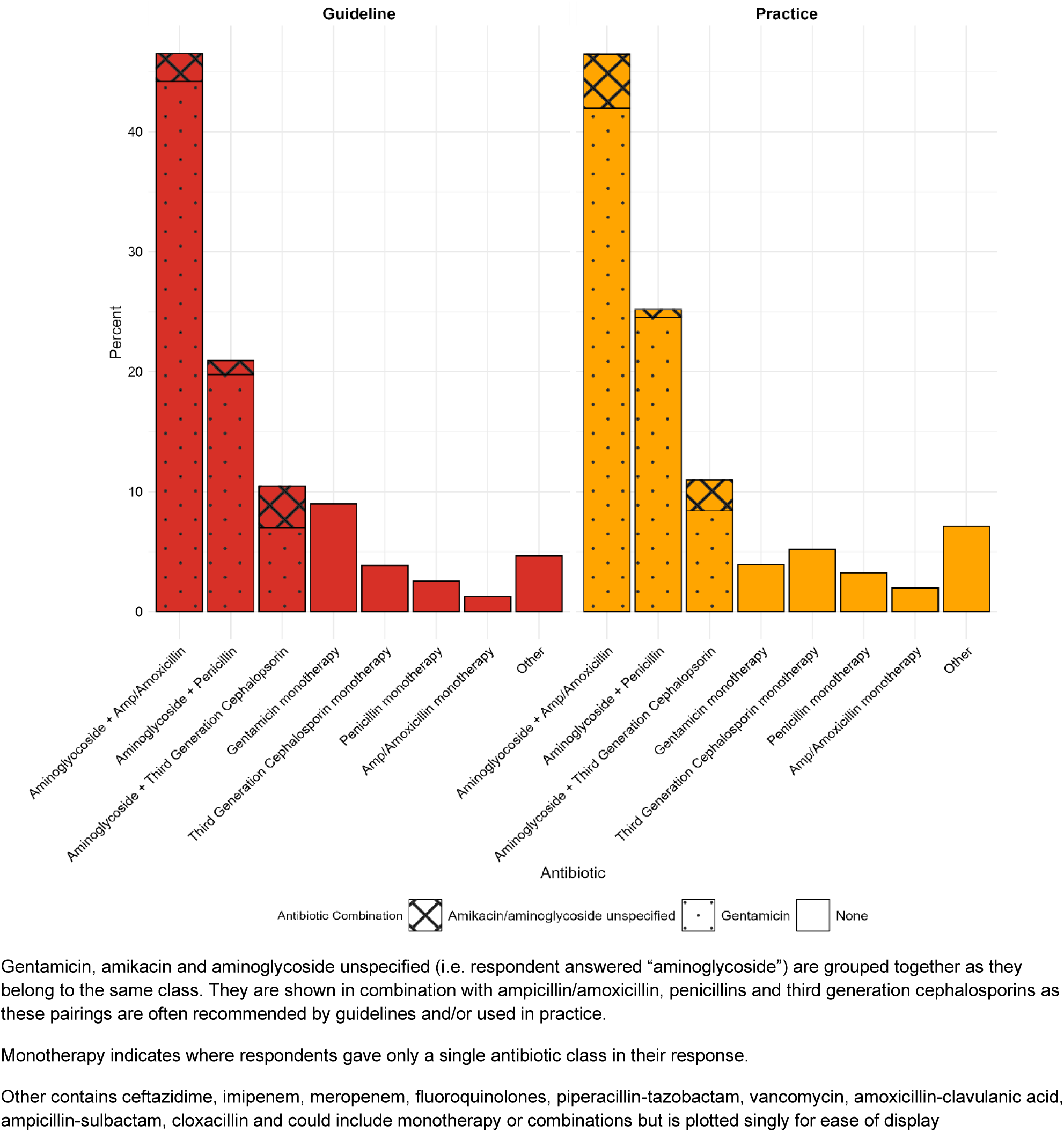
Antibiotics Used and Advised by Guidelines for Treating Early Onset Neonatal Sepsis.

Guidelines were reviewed in 37.7% of respondents’ settings at least every two years, with a further 20.8% not sure, and 41.5% who did not have their guidelines reviewed biannually. 45.4% had attempted to or been involved in the authorship of local guidelines during their careers. Of those who had not done so, the most common barrier that they faced was a lack of local AMR data (45.6% of responses). Respondents also reported lack of time (28.9%), cost of antibiotics (28.9%), lack of senior support (27.8%) and lack of experience (26.7%) as barriers to local guideline authorship.

## Discussion

This study presents the most comprehensive cross-sectional analysis to date of neonatal sepsis management practices in SSA, and substantiates concerns raised by smaller-scale studies, anecdotal work and expert opinion[27]. Our findings confirm widespread challenges in accessing timely microbiological diagnostic testing and appropriate antibiotic agents enabling optimal neonatal sepsis care. This study highlights almost universal utilisation and reported adherence to guidelines in practice. Across countries, guidelines commonly mirror WHO first-line antibiotic advice. This is despite current AMR patterns which demonstrate significant AMR to many of the antibiotics available and recommended in these guidelines.

Our study emphasises the scale of the diagnostic access challenge across SSA, highlighting both difficulties with access to diagnostics and timeliness of results to enable them to be clinically meaningful. Less than one quarter of clinicians were always able to obtain timely blood culture results to support their clinical decision-making and timely diagnostics for neonatal sepsis were only available half the time or less for cerebrospinal fluid, blood, urine and respiratory cultures. This echoes findings in other laboratory specialities in SSA that even when tests are available, turnaround times are often too long to impact patient care[28, 29]. With our respondents largely based in central hospitals, even this is likely an over-estimate, given that central healthcare facilities have greater diagnostic capacity than district and primary-level sites in SSA[30, 31]. Considering income, respondents from low- and LMIC settings had very similar challenges, and only those in five upper-middle income countries (Botswana, Gabon, Mauritius, Namibia, South Africa) had timely results more routinely available, suggesting that moderate economic growth does not necessarily translate into improved diagnostic capacity. Nevertheless, the introduction of rapid diagnostics for tuberculosis presents a hugely successful precedent for SSA countries, with reductions in turnaround times and improvements in diagnosis, access to treatment, cost-efficiency and patient outcomes[32].

Clinicians regularly faced challenges accessing antibiotics used to treat neonatal sepsis. First-line agents, such as ampicillin or amoxicillin, other penicillins, and gentamicin were most available, although even these were inconsistently accessible. Critically, national-level studies across SSA consistently demonstrate a high prevalence of multi-drug-resistant Gram-negative organisms causing neonatal sepsis[33, 34], often rendering first-line regimens ineffective. While first-line agents are cheap and available, effective escalation options providing adequate organism cover, such as carbapenems[35], as well as piperacillin-tazobactam were least available in our study. This is consistent with previous studies highlighting the lack of access and the prohibitive expense of escalation options despite these antibiotics’ necessity in LMICs[36]. Although the predominant use of WHO AWaRe ‘Access’ antibiotics may be interpreted as success[26] where available, ‘Watch’ and ‘Reserve antibiotics are widely used in SSA[37]. Therefore, our findings likely reflect limited availability of antibiotics rather than optimal antimicrobial stewardship, prompting calls to ensure that clinical need is also considered and drives access to appropriate antibiotics, including escalation options, when no other effective therapy is available[38]. Decisions on when and which antibiotics to start are predominantly based on guidelines, alongside neonates’ clinical condition. Our study indicates that guidelines are almost universally consulted, a previously unexplored yet key factor in understanding clinical decision-making surrounding antibiotic usage in neonatal sepsis in SSA. Local, national and WHO guidelines are all widely used, although national policies often mirror WHO advice[39], despite growing concerns that Africa-specific pathogen/AMR data and other key contextual factors may be missing[40].

Combination therapy with penicillins and gentamicin was the first-line antibiotic regimen used by most respondents, in guidelines and practice, aligning with current WHO guidance[41]. This is consistent with findings from a smaller survey by Rose-Mangeret *et al*.[11], which found this WHO-based approach to be most common in SSA, followed by using third-generation cephalosporins in combination with aminoglycosides. However, the NeoOBS cohort study, which investigated prescribing in neonatal sepsis across 11 LMICs, including three in Africa, found that only 25.9% started WHO first-line therapy and a further 13.8% cephalosporins[42]. While cephalosporin use in our cohort was similar, the use of first-line therapy was markedly higher in our cohort compared to theirs. This discrepancy may reflect our exclusive focus on Africa whilst NeoOBS included LMIC settings in South America and Asia. NeoOBS also centred on tertiary academic centres with better access to a wider array of antibiotic agents. It also observed practice, whilst we present self-reported data, in which desirability bias is possible. Understanding that the WHO-advised first-line regimens remain the predominant treatment approach in SSA is a critical finding of our work. This is especially relevant given the high prevalence of AMR amongst organisms causing neonatal sepsis towards first-line recommended antimicrobials in WHO guidelines[17–20].

Despite our cohort comprising largely senior doctors, fewer than half had participated in authoring antibiotic guidelines. In addition, we found that regardless of which guidelines were used, a significant proportion of settings did not routinely review or update them, which is critical to ensuring relevance and applicability to clinical practice[43]. The development of context-appropriate guidelines was primarily hindered by limited local AMR data, due to challenges in diagnostic capacity and surveillance. Together with limited antibiotic access and affordability, and lack of institutional support for developing guidelines, challenges reflect structural, political and resource constraints outlined previously[44]. Where implemented, guidelines can integrate local AMR data and diagnostic capability to inform antibiotic prescribing, making treatment more contextually relevant[45]. Given their widespread use among our respondents, guidelines can be powerful tools to drive targeted antimicrobial use, reduce unnecessary consumption and ultimately improve patient outcomes[46]. Conversely, a lack of locally relevant guidelines may undermine these objectives[47]. Context-specific guidelines can also prompt policy changes, such as to national essential medicine lists, to align antibiotic access with local AMR patterns[48]. Empowering local guideline development is therefore crucial to address rising, regionally specific AMR. Building on the WHO Global Action Plan for AMR, this includes aiming for improved access to diagnostics and antibiotics, empowerment of clinicians and improved data sharing between sites[43].

The number of respondents and breadth of country coverage are key strengths of our work, surpassing any previously published survey on neonatal sepsis in SSA. We explore the spectrum from investigation to management of neonatal sepsis, encompassing diagnostic access, prescribing practices and the development of guidelines. Although this survey represents a huge geographic region, for which solutions will not be uniform, this offers great perspective on SSA, often underserved by large-scale work, and addresses research gaps highlighted through previous work in neonatal sepsis. Study limitations include our presentation of reported, rather than observed practice. Given a recent study in two SSA countries highlighting differences between practice and guideline recommendations in neonatal sepsis[49] this approach leaves the possibility of desirability bias, although we aimed to mitigate this through survey anonymity. This study did not explore the mechanisms through which antibiotics are obtained by hospitals and clinicians, which may represent an important driver of AMR given the documented circulation of sub-standard and falsified antibiotics in SSA. [50]. Our respondents largely represent senior doctors, working in central healthcare facilities, reflecting our dissemination approach. However, neonates with sepsis may present to any level of care facility[27] and receive care provided by nurses, other healthcare workers or junior clinicians, who were not well represented in our study and who tend to dominate the workforce of primary and secondary facilities, particularly in rural areas[51].

Future research should deepen our understanding of whether poor outcomes from neonatal sepsis in SSA are driven by the mismatch between commonly used antibiotics and the heavy burden of AMR, as well as limitations in diagnostic capacity and data sharing. Innovative approaches to these issues, developed specifically for SSA must be trialled and promoted, regionally and globally. Qualitative research is also essential to explore the reasons underpinning reported behaviours, in utilisation of antibiotics, diagnostics and guidelines. Crucially, this must be seen as a global health priority, building on previous successes in other infectious diseases to drive collective action ensuring access to affordable, high-quality diagnostics and appropriate antibiotics to improve neonatal outcomes in SSA.

## List of Abbreviations

AMR: Antimicrobial resistance
LMIC: Low- and middle-income countries
SDG: Sustainable Development Goal
SSA: sub-Saharan Africa
WHO: World Health Organization

## Declarations

### Ethics approval and consent to participate

This study was approved by the Stellenbosch University Health Research Ethics Committee (HREC approval number: N24/10/118).

Informed consent was provided by each participant at the start of the survey having read the study’s Participant Information Sheet.

### Consent for publication

Not applicable.

### Availability of data and materials

As our data cover multiple countries with differing data protection laws, some of which are much more conservative about data sharing, even if anonymised, we cannot currently provide an open-source anonymised dataset. Further, there is risk of deductive disclosure in the several circumstances of small numbers of respondents per country.

### Authors’ contributions

J.S., D.H., R.P., L.B., G.C., F.F. and A.D. conceptualised the study and developed the initial survey tool. E.G. and I.C. translated the survey tool into French and Portuguese respectively. All authors contributed to survey dissemination via the networks described in the manuscript.

D.H., L.B. and A.D. managed data collection via REDCap. J.S., D.H. and L.B. curated and cleaned the data. J.S. and D.H. conducted data analysis. V.L., M.A.F., F.F. and A.D. supervised the work. J.S., D.H. and R.P. drafted the original manuscript. All authors provided critical feedback, read and approved the final manuscript.

### Acknowledgements

Not applicable.

## Supporting information

Supplementary Material - Survey

## Data Availability

All data produced in the present study are available upon reasonable request to the authors

---

**Supplementary Figure 1.**
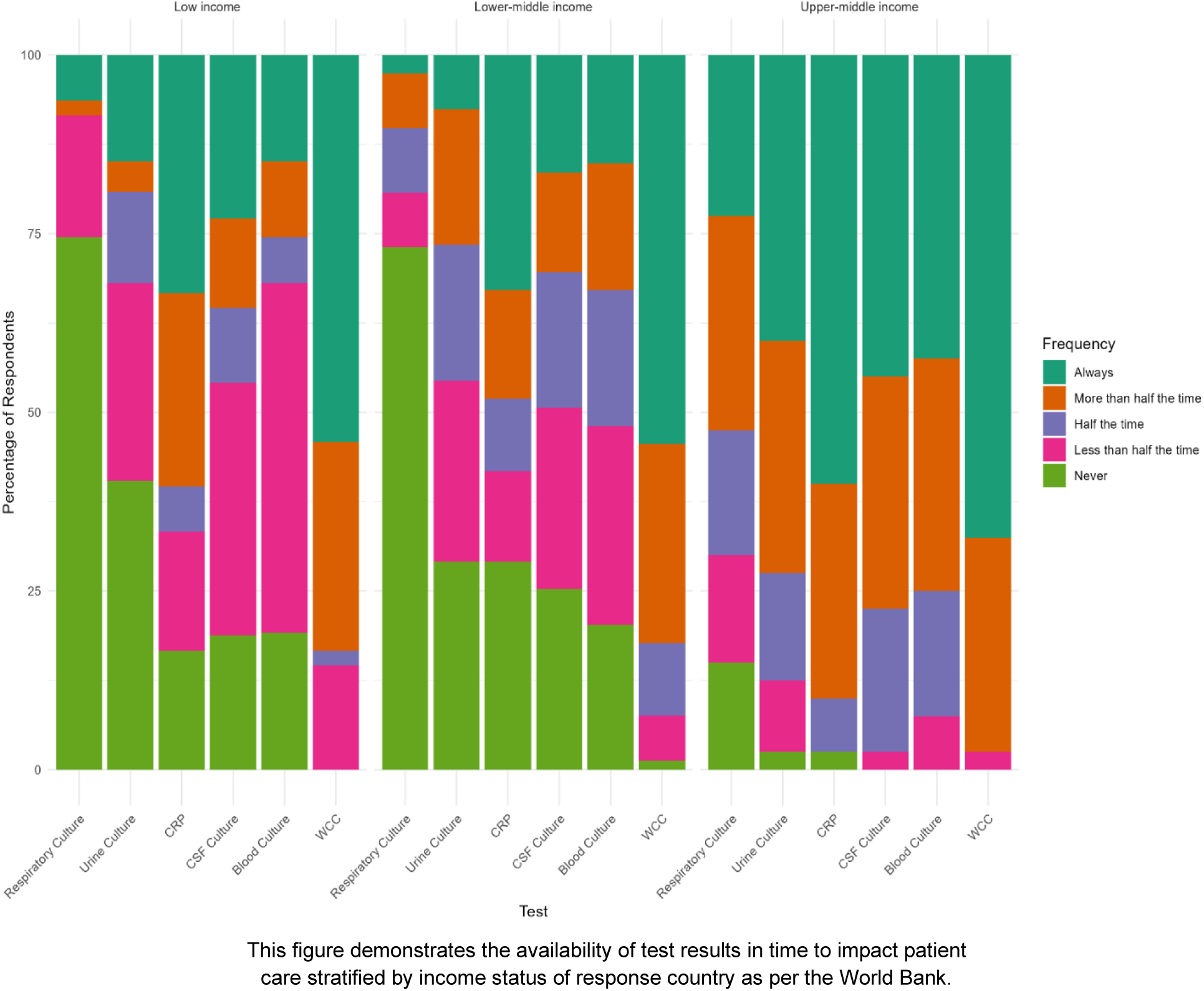
Timely Test Access by Location.

**Supplementary Figure 2.**
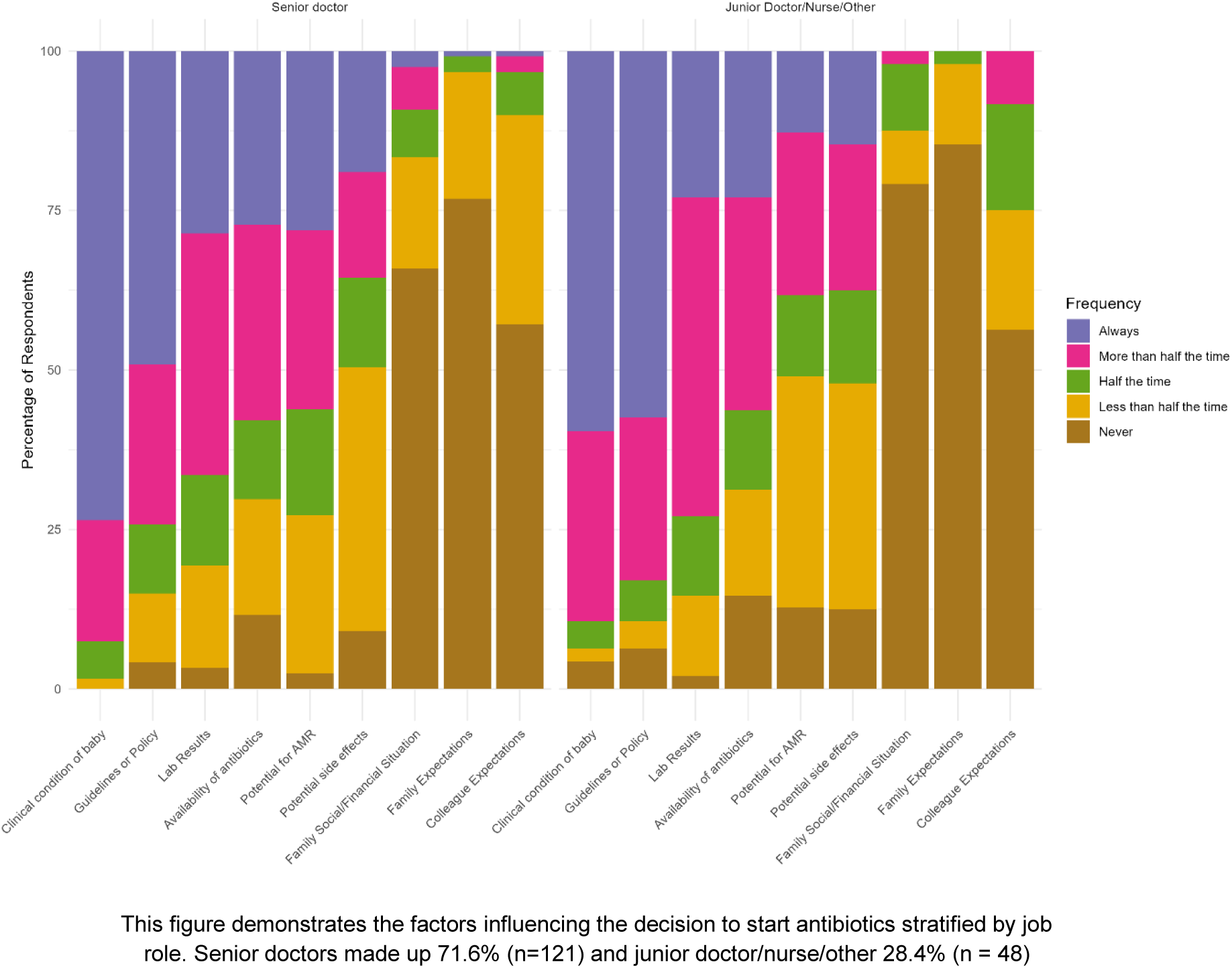
Influences on Decision to Start Antibiotics by Job Role.

**Supplementary Figure 3.**
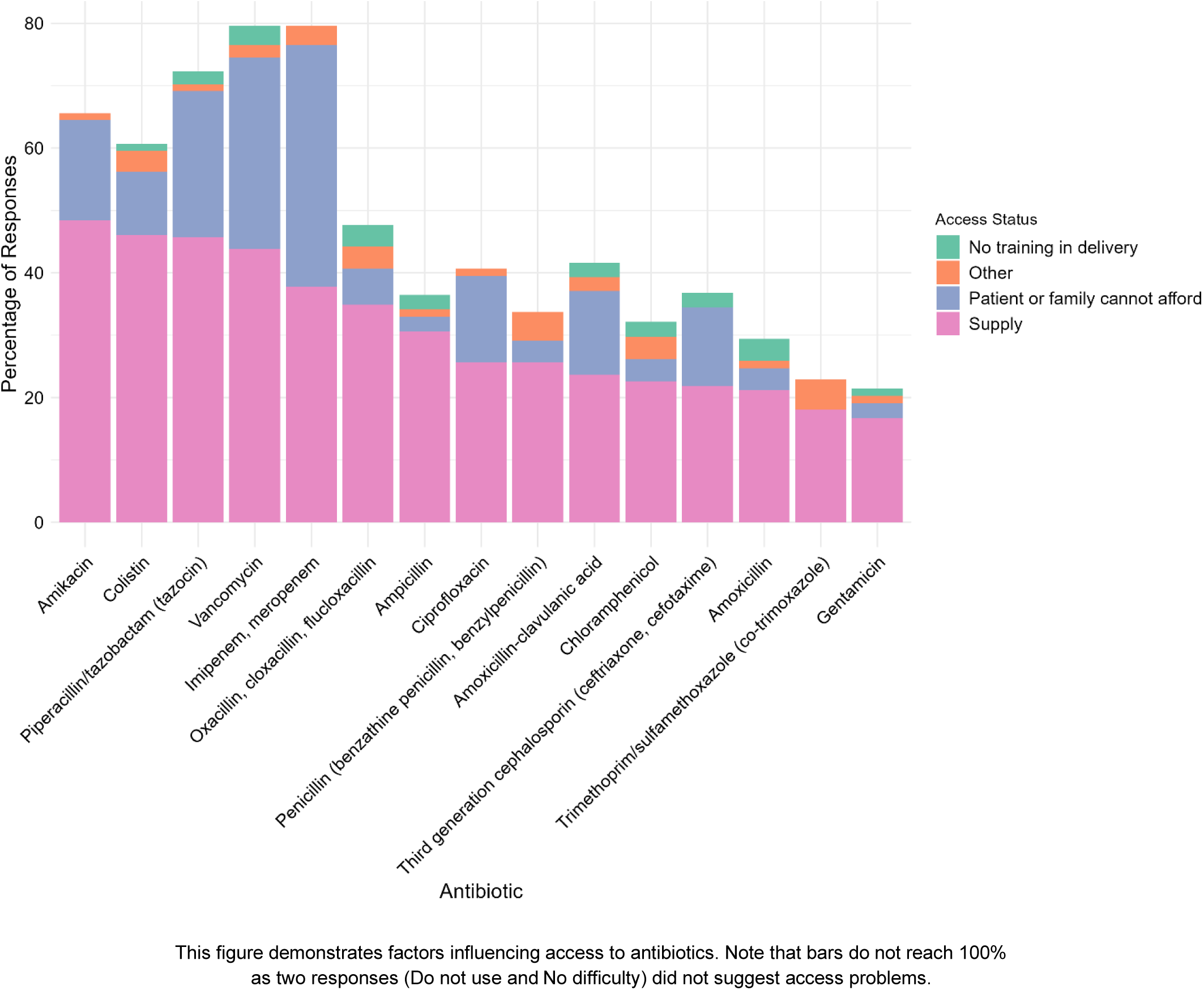
Factors Influencing Antibiotic Access.

**Supplementary Material – Survey Tool**

