## Supplementary Material - Survey for "Investigating Neonatal Sepsis: Anti-Infectives, Diagnostics and Guidelines used in Health sysTems across Sub-Saharan Africa - The INSIGHTS Study"

### Exploring practices in diagnostics, antimicrobials and guideline utilisation alongside establishing a minimum surveillance dataset for the management of neonatal sepsis in sub-Saharan Africa

#### Demographics

| | | |
|---|---|---|
| 1 | What is your job role? | ◯ Senior doctor (Neonatal clinician, paediatrician, consultant)<br>◯ Junior doctor (Intern, medical officer, registrar)<br>◯ Nurse (neonatal/paediatric)<br>◯ Other |
| 1a | Please specify | __________________________________ |
| 2 | For how many years have you practised professionally, after graduation? | ◯ < 5 years<br>◯ 5-10 years<br>◯ 11-20 years<br>◯ >20 years |
| 3 | Please specify which country you practise in. | __________________________________ |

| | | |
|---|---|---|
| 4 | In what type of healthcare facility do you work? | ◯ Public<br>◯ Private<br>◯ Both public and private<br>◯ Other |
| 4a | Please specify | ______________________________ |
| 5 | In which healthcare place do you practice? (tick all which apply) | ☐ Primary healthcare facility (e.g., hospital or clinic)<br>☐ District level (secondary level) hospital/healthcare facility<br>☐ Central (tertiary or quaternary) hospital/healthcare facility<br>☐ Laboratory<br>☐ Academic/Research institution (including staff involved in clinical duties)<br>☐ Other |
| 5a | Please specify | ______________________________ |
| 6 | What is the average number of neonatal inpatients at your hospital/healthcare facility at any given time? | ◯ < 20<br>◯ 21-40<br>◯ 41-60<br>◯ 61-100<br>◯ >100 |
| 7 | How often do you manage patients with neonatal sepsis? | ◯ Daily<br>◯ Weekly<br>◯ Monthly<br>◯ Yearly<br>◯ Less than yearly |

Diagnostics in neonatal sepsis

**8. Do you have access to the below diagnostic services to support you in the care of patients with neonatal sepsis?**

| | Never | Less than half of the time | Half of the time | More than half of the time | Always |
|---|---|---|---|---|---|
| Blood white cell count | ◯ | ◯ | ◯ | ◯ | ◯ |
| C-reactive protein | ◯ | ◯ | ◯ | ◯ | ◯ |
| Blood culture | ◯ | ◯ | ◯ | ◯ | ◯ |
| Urine culture | ◯ | ◯ | ◯ | ◯ | ◯ |
| Respiratory (bacterial) culture | ◯ | ◯ | ◯ | ◯ | ◯ |
| Cerebrospinal fluid (CSF) culture | ◯ | ◯ | ◯ | ◯ | ◯ |

#### 9. How often in the last month have you ordered/performed each test?

| | Never | 1-5 | 6-10 | 11-20 | 21-50 | 51-100 | >100 |
|---|---|---|---|---|---|---|---|
| Blood white cell count | ○ | ○ | ○ | ○ | ○ | ○ | ○ |
| C-reactive protein | ○ | ○ | ○ | ○ | ○ | ○ | ○ |
| Blood culture | ○ | ○ | ○ | ○ | ○ | ○ | ○ |
| Urine culture | ○ | ○ | ○ | ○ | ○ | ○ | ○ |
| Respiratory (bacterial) culture | ○ | ○ | ○ | ○ | ○ | ○ | ○ |
| Cerebrospinal fluid (CSF) culture | ○ | ○ | ○ | ○ | ○ | ○ | ○ |

#### 10. How often is the result of this test available to you quickly enough to impact your patients' care?

| | Never | Less than half of the time | Half of the time | More than half of the time | Always |
|---|---|---|---|---|---|
| Blood white cell count | ○ | ○ | ○ | ○ | ○ |
| C-reactive protein | ○ | ○ | ○ | ○ | ○ |
| Blood culture | ○ | ○ | ○ | ○ | ○ |
| Urine culture | ○ | ○ | ○ | ○ | ○ |
| Respiratory (bacterial) culture | ○ | ○ | ○ | ○ | ○ |
| Cerebrospinal fluid (CSF) culture | ○ | ○ | ○ | ○ | ○ |

#### 11. How often do you have difficulty with each of the following in terms of microbiology services?

| | Never | Less than half of the time | Half of the time | More than half of the time | Always |
|---|---|---|---|---|---|
| No laboratory service available | ○ | ○ | ○ | ○ | ○ |
| Lack of consumables to perform tests from the clinical side (e.g. needles, blood culture bottles, lumbar puncture materials) | ○ | ○ | ○ | ○ | ○ |
| Transport/delivery of samples to the laboratory | ○ | ○ | ○ | ○ | ○ |
| Samples not being accepted by the laboratory | ○ | ○ | ○ | ○ | ○ |
| Patients being unable to afford recommended tests | ○ | ○ | ○ | ○ | ○ |
| Tests not being completed by the laboratory | ○ | ○ | ○ | ○ | ○ |
| Laboratory results do not return to me | ○ | ○ | ○ | ○ | ○ |
| Laboratory results received too late to impact patient care | ○ | ○ | ○ | ○ | ○ |
| Laboratory results are not reliable | ○ | ○ | ○ | ○ | ○ |

| | | | | | |
|---|---|---|---|---|---|
| Laboratory results are from samples taken after antibiotics have already been administered? | ○ | ○ | ○ | ○ | ○ |
| Other | ○ | ○ | ○ | ○ | ○ |

| | Never | Less than half of the time | Half of the time | More than half of the time | Always | I don't know about this issue |
|---|---|---|---|---|---|---|
| Lack of consumables to perform tests from the laboratory side (e.g. antibiotic disks, agars, slides) | ○ | ○ | ○ | ○ | ○ | ○ |
| High workload in the laboratory (sample numbers or staff shortages) | ○ | ○ | ○ | ○ | ○ | ○ |

11a If you selected anything other than 'Never' for other, please specify the issue. __________________________________

Antibiotic management in neonatal sepsis

**12. Which of these factors influence your decision whether to start antibiotic treatment or not in a neonate?**

| | Never | Less than half of the time | Half of the time | More than half of the time | Always |
|---|---|---|---|---|---|
| Clinical condition of the baby | ○ | ○ | ○ | ○ | ○ |
| Expectations of the family | ○ | ○ | ○ | ○ | ○ |
| Expectations of colleagues | ○ | ○ | ○ | ○ | ○ |
| Social/financial situation of the family | ○ | ○ | ○ | ○ | ○ |
| Guidelines/policy | ○ | ○ | ○ | ○ | ○ |
| Laboratory results | ○ | ○ | ○ | ○ | ○ |
| Potential development of antimicrobial resistance | ○ | ○ | ○ | ○ | ○ |
| Availability of any appropriate antibiotics | ○ | ○ | ○ | ○ | ○ |
| Potential side effects of antibiotics | ○ | ○ | ○ | ○ | ○ |

**13. Once you have made the decision to start antibiotics, what influences your decision on which antibiotic(s) to prescribe in neonatal sepsis?**

| | Yes | No |
|---|---|---|
| Personal experience/training | ○ | ○ |
| Advice of neonatal clinicians | ○ | ○ |
| Advice of microbiologists | ○ | ○ |

| | | |
|---|---|---|
| Local guidelines | ◯ | ◯ |
| National guidelines | ◯ | ◯ |
| International guidelines | ◯ | ◯ |
| Clinical condition of the baby | ◯ | ◯ |
| Cost of antibiotics | ◯ | ◯ |
| Availability of antibiotics | ◯ | ◯ |
| Other | ◯ | ◯ |

13a Please specify

__________________________________

14 Do you have any challenges accessing antibiotics in your practice managing neonatal sepsis?

◯ Yes
◯ No

14a How often do you have difficulty accessing each of the following antibiotics to use in patients with neonatal sepsis?

| | Never | Less than half of the time | Half of the time | More than half of the time | Always | Would not use this antibiotic |
|---|---|---|---|---|---|---|
| Penicillin (benzathine penicillin, benzylpenicillin) | ◯ | ◯ | ◯ | ◯ | ◯ | ◯ |
| Ampicillin | ◯ | ◯ | ◯ | ◯ | ◯ | ◯ |
| Amoxicillin | ◯ | ◯ | ◯ | ◯ | ◯ | ◯ |
| Oxacillin, cloxacillin, flucloxacillin | ◯ | ◯ | ◯ | ◯ | ◯ | ◯ |
| Ceftriaxone, cefotaxime (3rd generation cephalosporin) | ◯ | ◯ | ◯ | ◯ | ◯ | ◯ |
| Gentamicin | ◯ | ◯ | ◯ | ◯ | ◯ | ◯ |
| Amikacin | ◯ | ◯ | ◯ | ◯ | ◯ | ◯ |
| Imipenem, meropenem | ◯ | ◯ | ◯ | ◯ | ◯ | ◯ |
| Ciprofloxacin | ◯ | ◯ | ◯ | ◯ | ◯ | ◯ |
| Azithromycin, erythromycin | ◯ | ◯ | ◯ | ◯ | ◯ | ◯ |
| Chloramphenicol | ◯ | ◯ | ◯ | ◯ | ◯ | ◯ |
| Co-trimoxazole | ◯ | ◯ | ◯ | ◯ | ◯ | ◯ |
| Vancomycin | ◯ | ◯ | ◯ | ◯ | ◯ | ◯ |
| Amoxicillin/clavulanic acid (co-amoxiclav) | ◯ | ◯ | ◯ | ◯ | ◯ | ◯ |
| Piperacillin/tazobactam (tazocin) | ◯ | ◯ | ◯ | ◯ | ◯ | ◯ |
| Colistin | ◯ | ◯ | ◯ | ◯ | ◯ | ◯ |
| Fluconazole | ◯ | ◯ | ◯ | ◯ | ◯ | ◯ |
| Amphotericin B | ◯ | ◯ | ◯ | ◯ | ◯ | ◯ |
| Aciclovir (acyclovir) | ◯ | ◯ | ◯ | ◯ | ◯ | ◯ |

14b What are the difficulties that you have in using these antibiotics?

| | Out of stock antibiotic/no supply to healthcare facility | Patient/family unable to afford medications | Inadequate training to deliver the medication | Other | Do not use this antibiotic in practice | No difficulty with access to this antibiotic |
|---|---|---|---|---|---|---|
| Penicillin (benzathine penicillin, benzylpenicillin) | ☐ | ☐ | ☐ | ☐ | ☐ | ☐ |
| Ampicillin | ☐ | ☐ | ☐ | ☐ | ☐ | ☐ |
| Amoxicillin | ☐ | ☐ | ☐ | ☐ | ☐ | ☐ |
| Oxacillin, cloxacillin, flucloxacillin | ☐ | ☐ | ☐ | ☐ | ☐ | ☐ |
| Ceftriaxone, cefotaxime (3rd generation cephalosporin) | ☐ | ☐ | ☐ | ☐ | ☐ | ☐ |
| Gentamicin | ☐ | ☐ | ☐ | ☐ | ☐ | ☐ |
| Amikacin | ☐ | ☐ | ☐ | ☐ | ☐ | ☐ |
| Imipenem, meropenem | ☐ | ☐ | ☐ | ☐ | ☐ | ☐ |
| Ciprofloxacin | ☐ | ☐ | ☐ | ☐ | ☐ | ☐ |
| Azithromycin, erythromycin | ☐ | ☐ | ☐ | ☐ | ☐ | ☐ |
| Chloramphenicol | ☐ | ☐ | ☐ | ☐ | ☐ | ☐ |
| Co-trimoxazole | ☐ | ☐ | ☐ | ☐ | ☐ | ☐ |
| Vancomycin | ☐ | ☐ | ☐ | ☐ | ☐ | ☐ |
| Amoxicillin/clavulanic acid (co-amoxiclav) | ☐ | ☐ | ☐ | ☐ | ☐ | ☐ |
| Piperacillin/tazobactam (tazocin) | ☐ | ☐ | ☐ | ☐ | ☐ | ☐ |
| Colistin | ☐ | ☐ | ☐ | ☐ | ☐ | ☐ |
| Fluconazole | ☐ | ☐ | ☐ | ☐ | ☐ | ☐ |
| Amphotericin B | ☐ | ☐ | ☐ | ☐ | ☐ | ☐ |
| Aciclovir (acyclovir) | ☐ | ☐ | ☐ | ☐ | ☐ | ☐ |

14b-iPlease specify

__________________________________

| | | |
|---|---|---|
| 15 | Do you have pharmacy support available in managing your antibiotic decisions? | ◯ Never<br>◯ Less than half of the time<br>◯ Half of the time<br>◯ More than half of the time<br>◯ Always<br>◯ Would not use/consult pharmacy for this |
| 16 | Is there an antimicrobial stewardship (AMS) team/action plan specific to your facility? | ◯ Yes<br>◯ No<br>◯ Not sure<br>(This would not be in the form of a National Action Plan, it must be facility specific) |
| 17 | Are there regular audits or reviews of the use of antibiotics in your facility? | ◯ Yes<br>◯ No<br>◯ Not sure |

#### The use of guidelines in neonatal sepsis

| | | |
|---|---|---|
| 18 | Do you refer to treatment guidelines in the management of neonatal sepsis? | ◯ Yes<br>◯ No |
| 19 | Which, if any, antibiotic guideline is used for treating neonatal sepsis at your healthcare facility? | ☐ Yes - local guideline (specific to my healthcare facility)<br>☐ Yes - national guideline<br>☐ Yes - WHO guideline<br>☐ Other guideline<br>☐ No guideline |
| 19a | Please specify which guideline | __________________________________ |
| 19b | Do you find these antibiotic guidelines applicable to your setting? | ◯ Yes<br>◯ No<br>◯ Sometimes |
| 19b-i | Please describe why the antibiotic guidelines are not always applicable to your practice? | __________________________________ |
| 19c | Are the antibiotic guidelines used at your healthcare facility regularly reviewed and updated considering new evidence (at least every 2 years)? | ◯ Yes<br>◯ No<br>◯ Not sure |
| 20 | Have you or your colleagues attempted to author any local or national specific antibiotic guidelines? | ◯ Yes<br>◯ No |
| 20a | What challenges or support did you encounter during this process? | __________________________________ |

20b What factors have influenced this?

- ☐ Not enough time to develop
- ☐ Do not feel these are needed
- ☐ Lack of support from colleagues
- ☐ Lack of support from senior hospital staff e.g. medical superintendent, clinical director
- ☐ Financial constraints e.g. could not afford alternative antibiotics to those currently used
- ☐ Availability of other antibiotic agents/options
- ☐ Lack of data on local antimicrobial resistance patterns to guide local guideline development
- ☐ I do not feel experienced enough to contribute towards local guideline development
- ☐ Other

20b-i Please specify which other factors have influenced this

__________________________________

21 When treating EARLY ONSET neonatal sepsis at your facility what empiric first line antibiotic regimen:

a. do you use in clinical practice?

__________________________________
(Please include agent(s) and route of administration (i.e. IV or oral))

21 b. is recommended by your guideline?

__________________________________
(Please include agent(s) and route of administration (i.e. IV or oral))

22 When treating LATE ONSET HOSPITAL-ACQUIRED neonatal sepsis at your facility what empiric first line antibiotic regimen:

a. do you use in clinical practice?

__________________________________
(Please include agent(s) and route of administration (i.e. IV or oral))

22 b. is recommended by your guideline

__________________________________
(Please include agent(s) and route of administration (i.e. IV or oral))

23 When treating LATE ONSET COMMUNITY-ACQUIRED neonatal sepsis at your facility what empiric first line antibiotic regimen:

a. do you use in clinical practice?

__________________________________
(Please include agent(s) and route of administration (i.e. IV or oral))

23 b. is recommended by your guideline?

__________________________________
(Please include agent(s) and route of administration (i.e. IV or oral))

24 How often are you able to adhere to the antibiotic guidelines at your facility?

- ◯ Never
- ◯ Very rarely
- ◯ Less than half of the time
- ◯ Half of the time
- ◯ More than half of the time
- ◯ Always

25 If you divert from the antibiotic guidelines, why do you do this?

- ☐ Clinical presentation of the neonate
- ☐ Opinion/concern of colleagues on the condition of the neonate
- ☐ Unavailabilty of the antibiotics suggested in the guideline
- ☐ Aware of antibiotic-resistant organisms not addressed by the guideline
- ☐ Availability of equipment (e.g. cannula) to deliver antibiotic suggested in the guideline
- ☐ No intravenous access for antibiotic delivery
- ☐ Financial considerations for family
- ☐ Never have to divert from antibiotic guidelines
- ☐ Other

25a Please specify

__________________________________

26 Do you face any barriers in diverting from antibiotic guidelines if you feel it appropriate to do so?

- ◯ Yes
- ◯ No

26a Describe the issues which you face when diverting from antibiotic guidelines.

__________________________________________

Surveillance in neonatal sepsis

**27. Does your facility collect surveillance data on...?**

| | Yes | No |
|---|---|---|
| Patients with suspected neonatal infection | ◯ | ◯ |
| Patients with confirmed neonatal bloodstream infection or meningitis | ◯ | ◯ |
| Neonatal mortality (crude) | ◯ | ◯ |
| Neonatal mortality from sepsis | ◯ | ◯ |
| Antibiotic use in the neonatal unit | ◯ | ◯ |
| No aspects of neonatal sepsis | ◯ | ◯ |

27a Who collects surveillance data?

- ☐ Clinicians
- ☐ Hospital records team
- ☐ Pharmacists
- ☐ Public Health
- ☐ Microbiology laboratory
- ☐ National Health Information System
- ☐ Other

27a-i Please specify

__________________________________

#### Comments

28 Do you have any further comments?

_______________________________
